# Impact of a low-threshold exercise and protein supplementation intervention to optimise physical function and target frailty in people experiencing homelessness and addiction: The LEAP II trial

**DOI:** 10.1101/2024.10.27.24316246

**Authors:** Fiona Kennedy, Clíona Ní Cheallaigh, Roman Romero-Ortuno, Deirdre Murray, Julie Broderick

## Abstract

**Objective:** Frailty is a state of physiological vulnerability and associated with adverse outcomes. People experiencing homelessness and addiction have a higher burden of frailty than the general population. This study’s aim was to test the feasibility and impact of an exercise intervention with protein supplementation to target low physical functioning and frailty in this population, which was flexibly delivered over 12 weeks, with exercise options three days per week.

**Methods:** The primary outcome of feasibility was measured by recruitment, retention, adherence, acceptability and adverse events. Secondary outcomes were physical function, pain, frailty and nutritional status, and self-reported health.

**Results:** Forty-three participants enrolled. Overall retention was 69.8%, with higher retention observed in subgroups (women and older adults). Programme adherence and acceptability was reported in 93% and 100% of participants, respectively. No adverse events occurred. At baseline, sub-normative values were demonstrated for limb strength and balance; pain was prevalent in 34.8%; 23% had poor nutritional health between 32.5% and 72.1% lived with some degree of frailty. Significant improvements were demonstrated for lower limb strength, gait speed, pain, nutritional status and frailty, (p<0.05).

**Conclusions:** This novel intervention delivered to this hard-to-reach population was shown to be feasible and impactful, indicating proof of concept.

## Introduction

Frailty is a complex state of cumulative decline which renders a person vulnerable to adverse health outcomes [1]. Frailty is associated with lower socioeconomic status (SES) and has been identified in populations with lower SES and multiple disadvantage [2, 3]. Poor physical functioning, premature ageing and high levels of frailty has been reported in people experiencing homelessness (PEH) [3, 4]. Unprecedented levels of homelessness are continually being reported globally and a record of 653,104 Americans experiencing homelessness was reported in 2023 [5]. People experiencing homelessness are at high risk of substance misuse. Moreover, an association between a tri-morbidity of substance misuse, mental and physical health conditions and PEH is commonly reported [6]. Substance use disorder (SUD) is a major public health concern and drug addiction is the leading risk factor for premature death globally [7]. This population also experience poor physical health and comorbid conditions including accidental injuries, lung or heart disease, diabetes, stroke and cancer [8, 9].

People experiencing homelessness and addiction face challenges accessing healthcare, which is mostly delivered in highly structured environments [10, 11]. Low-threshold, trauma informed interventions are advocated to address the complex needs identified and are shown to be effective in reducing substance use-related harm [12, 13]. Exercise has been proposed as an adjunctive therapy for SUD [14–17]. Much research on exercise in SUD has focused on substance use-related outcomes and a range of mental health outcomes [17, 18]. Despite the known poor physical health of PEH and addiction issues [3, 4, 19] there is a dearth of evidence on exercise interventions targeting physical function and frailty in this population [15, 18, 20].

Poor nutritional intake has been identified as a key driver of frailty [21]. Protein supplementation, in addition to exercise, particularly strength training, has proven effectiveness in reversing or delaying frailty [22, 23]. The impact of exercise with protein supplementation to target frailty in PEH was explored in the LEAP-I (Low threshold Exercise And Protein supplementation) trial [19]. While feasibility of the once-weekly intervention was demonstrated, recommendations for further exercise opportunities were made [19].

The aim of this study was to measure the feasibility and the impact of an enhanced low-threshold exercise intervention with multiple exercise opportunities each week with protein supplementation to target physical function and frailty in PEH and addiction.

## Methods

### Study design

This was a pre-post intervention study measuring the impact of a 12-week exercise programme with protein supplementation. The programme took place from October 3^rd^ 2022-March 3^rd^ 2023 in a day centre in Dublin, Ireland for people with chronic addiction issues, many of whom experienced homelessness. This study was approved by the Faculty of Health Sciences REC at Trinity College Dublin (Ethical Approval Reference Number: 211202).

A stakeholder engagement session was held in advance to build in a co-design approach with people with lived experience of addiction and homelessness. This focussed session informed the low-threshold and trauma informed design of the programme. Participant information leaflets were distributed via a gatekeeper in the centre. Eligibility screening, written informed consent and baseline assessment was initially conducted, and GPs were notified of participants involvement.

Any person (18-65 years) accessing services in the centre who consented to participate was included. Participants with problematic behavioural issues, major physical or cognitive impairments which precluded ability to safely participate or with a confirmed pregnancy were excluded.

### Outcomes

The primary outcome was feasibility measured by recruitment, retention, adherence, adverse events, and programme acceptability, as outline in Table 1.

**Table 1:** Primary Outcome.

| Primary Outcome:<br>Feasibility | Method of reporting |
| --- | --- |
| Recruitment Numbers | Number of eligible consenting participants who completed the initial assessment |
| Retention rate | Number of return visits and the frequency of attendance<br>-attending at least once/week for > 50% of the duration of the programme (regular attenders)<br>-attending at least once/week for < 50% of the duration of the programme (sporadic attenders)<br>-did not return following initial assessment (non-attenders).<br>-Attendance by gender |
| Sub-group retention rate | -Attendance by age |
| Adherence rate (to the exercise intervention) | Percentage of sessions adhered to/completed by participants |
| Adherence rate (to the nutritional supplement) | Percentage of participants who consumed the protein supplement drink |
| Adverse events | An unfavourable and unintended sign, symptom, or disease having been absent at baseline, or, if present at baseline, appears to worsen and is temporally associated with medical treatment or procedure, regardless of the attribution [24]. |
| Programme Acceptability | Participant feedback provided by Exit Survey; analysed by frequency of responses and observing patterns or main themes in the data collected. |

Secondary outcomes, measuring impact, were physical function and pain, frailty and nutritional status and self-reported health status, recorded at baseline and end of programme. Upper and lower limb strength were measured using handgrip dynamometry and the Chair Stand Test [25, 26, 27, 28]. Mid-calf and arm circumference measured muscle mass [29, 30]. Mid-arm circumference is recommended for use in PEH and addiction [4] due to the high prevalence of lower limb swelling [31]. The 10m Walk Test (10mWT) [32], the 2 Minute Walk Test (2MWT) [33, 34] and the Single Leg Stance Test (SLST) [35, 36] were conducted to measure gait speed, functional capacity and balance, respectively. Results were compared to normative reference values. Each participant was asked about pain, and severity was assessed using the Numerical Pain Rating Scale (NPRS) [37]. Frailty was explored using the Clinical Frailty Scale (CFS) [38] and SHARE-Frailty Instrument (SHARE-FI) [39]. Nutritional status was assessed using the Mini-nutritional assessment (MNA) [40, 41] and the Short-Form 12 (SF12) was used as a self-report evaluation of health [42, 43].

### Intervention

The 12-week intervention was designed to be low-threshold and participant-centred. Group, individual, and gender-based exercise classes were flexibly delivered by two research physiotherapists (FK and MM) over 15 weeks to facilitate maximum inclusion and participation. (*FK had over 25 years of clinical experience and MM was a staff grade physiotherapist*.) Classes were available in the mornings and afternoons on two alternate days and the Park Walk was held for one morning each week. The exercise classes were multi-modal with a primary focus on strength and were adapted and progressed based on the physical ability of the participants, and their often-fluctuating presentation and motivation (Table 2). Music was an important feature of the programme. Blood pressure and heart rate was monitored as a safety indicator and exercise intensity was low to moderate, and was modulated using Borg’s Perceived Rate of Exertion (RPE) scale, where participants were advised to exercise between 11 and 13 on the RPE scale [44]. To promote post-exercise muscle protein synthesis, a protein supplement (Fresubin, https://www.fresubin.com/) was offered to participants. Participants were educated about physical activity and provided with basic nutritional advice. They were encouraged to attend twice-weekly for the exercise classes and once-weekly for the ‘Park Walk’. Motivational phone calls were made to remind participants of the classes and address barriers to adherence.

**Table 2.** Exercise Circuit.

| Core exercise | Initial Intensity | Progression/Adaptations* |
| --- | --- | --- |
| Sit to stand or squats | 2 sets 10-15 reps | 3 sets of 15 repetitions. lunges, inner range quads; use of weights/ball |
| Elbow Bends | 2 sets 10-15 reps | 3 sets of 15 repetitions. use of weights |
| Step-ups | 2 sets 10-15 reps | 3 sets of 15 repetitions. alter height of step; use of weights |
| Arm elevations | 2 sets 10-15 reps | 3 sets of 15 repetitions. use of weights |
| Hip abductions | 2 sets 10-15 reps | 3 sets of 15 repetitions. with additional upper limb abduction and elevation; movement with 360° turns |
| Scapular retractions | 2 sets 10-15 reps | 3 sets of 15 repetitions. use of weights |
| Aerobic activity | 2 mins | 3-5 mins use of ladders, hurdles, skipping ropes, jumping jacks. dance, game with cones/balls |
| Balance | 4-5 mins | 5 mins tandem; single leg stance, upper limb, and trunk movements; weights and ball work |

Commenced with warm-up and ended with cool-down. \* Adaptations: exercises were individualised and progressed for each participant by the research physiotherapist Progression: 2-3 sets of 10-15 repetitions completed at each session; beyond that, low resistance weights (1.1kg-4.5kg), theraband or increased step height was introduced to achieve a strengthening effect.
Exercise equipment: therabands, dumbbells, steps, ladders, hurdles, skipping ropes and balls.

### Data analysis

Descriptive statistics were performed to summarise participant characteristics. Pre- and post-intervention data were compared using paired t-tests or Wilcoxon signed-rank tests. Data were analysed using IBM SPSS V28 and a *p*-value with statistical significance was set at *p*<0.05. Exit survey data were analysed by (i) observing frequencies of responses, (ii) exploring data for themes using a low level of interpretation and (iii) illustrating themes through verbatim comments, while bearing in mind that this represents ‘quasi-qualitative data’, not in-depth analysis addressed by qualitative study methodology [45].

## Results

### Feasibility

#### Recruitment

Forty-seven people were initially recruited. Forty-three participants progressed to study enrolment, 30 returned for the intervention and 20 completed the final evaluation (Fig 1).

**Figure 1.**
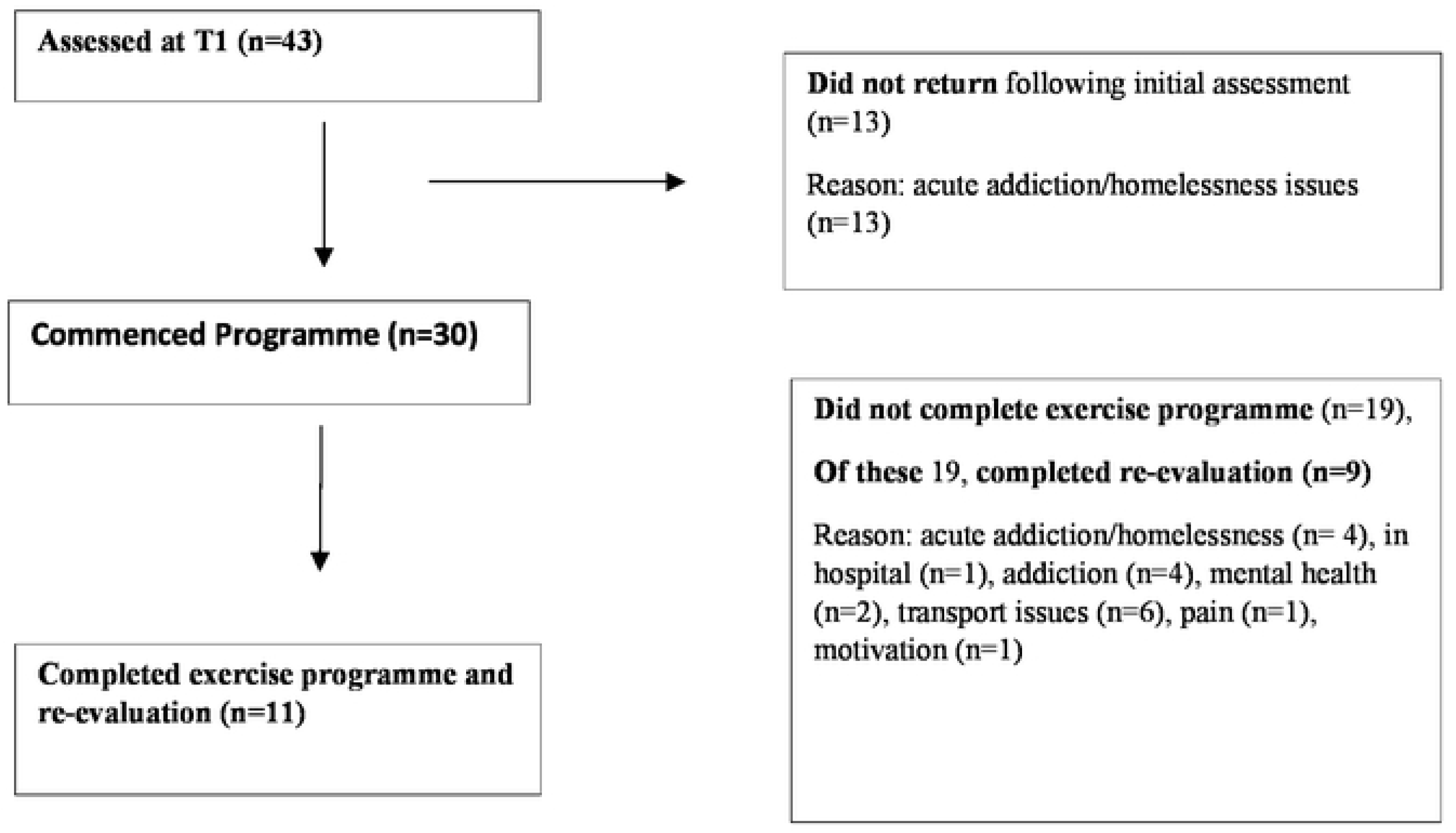
PRISMA Flow Diagram of participants through the study here.

The mean (SD) age was 45.6 (9.5) years. Just over half (53.5%, n=23) were female. Most were single (39.5%, n= 17) and unemployed (90.7%, n=39). Three-quarters (74.4%, n=32) were white Irish and 25.6% (n=11) were Irish Travellers. The majority (81.4%, n=35) experienced problematic substance use. Half (51.1%, n=22) of the participants were homeless. All participants had a self-reported mental health condition and were on prescribed medication for this. A range of physical health conditions were reported. Reminder phone calls of classes yielded a 0% return rate. Baseline data of participants including physical function and frailty, are presented in Table 3 and 4.

**Figure 2.**
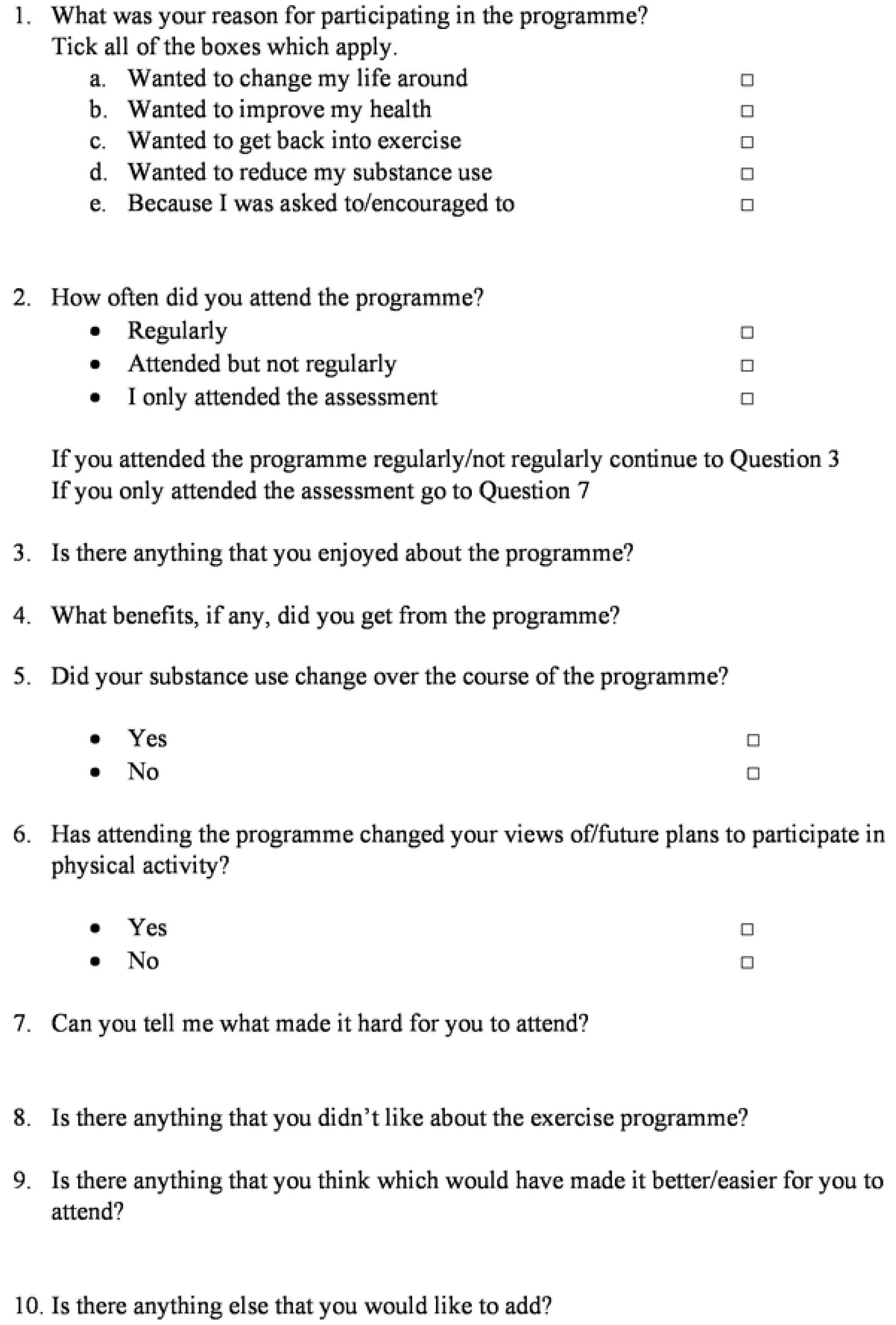
Exit Survey.

**Table 3.** Participant Demographics.

| **Demographic item** |  |
| --- | --- |
| Age mean (std dev) (range) | 45.6 (9.5) (26-62) years |
| Female n (%) | 23 (53.5) |
| Male n (%) | 20 (46.5) |
| **Marital status n (%)** |  |
| Single | 17 (39.5) |
| Married/partnership | 10 (23.3) |
| Separated/divorced | 8 (18.6) |
| Widowed | 7 (16.3) |
| *Data unavailable | 1 (2.3) |
| **Ethnicity n (%)** |  |
| White Irish | 32 (74.4) |
| Irish Traveller | 11 (25.6) |
| **Employment status n (%)** |  |
| Full-time employment | 1 (2.3) |
| Part-time employment | 3 (7) |
| Unemployed | 39 (90.7) |
| **\*\*Housing environment n (%)** |  |
| House/apartment | 21 (48.9) |
| Hotel/hostel/staying with friend/family/other | 8 (18.6) |
| Halting site | 9 (20.9) |
| Rough sleeper | 5 (11.6) |
| **Living Arrangement n (%)** |  |
| Alone | 18 (41.9) |
| With spouse/partner | 5 (11.6) |
| Other | 20 (46.5) |
| **History of Addiction n (%)** |  |
| Dependency | 27 (62.8) |
| Harmful to hazardous use | 4 (9.3) |
| Stable/in recovery | 4 (9.3) |
| No addiction issues | 8 (18.6) |
| **Physical health conditions n (%)** |  |
| Musculoskeletal | 18 (41.9) |
| Respiratory | 13 (30.2) |
| Cardiovascular | 11 (25.5) |
| Seizures | 6 (13.9) |
| Hepatitis | 6 (13.9) |
| Liver disease | 5 (11.6) |
| Diabetes | 4 (9.3) |
| Cancer | 3 (6.9) |
| HIV | 2 (4.6) |
| Other | 7 (16.3) |

\*some participants had >1 physical health condition
\*\*Homelessness status: aligned with the European Typology for Homelessness and Housing (ETHOS)

**Table 4.**
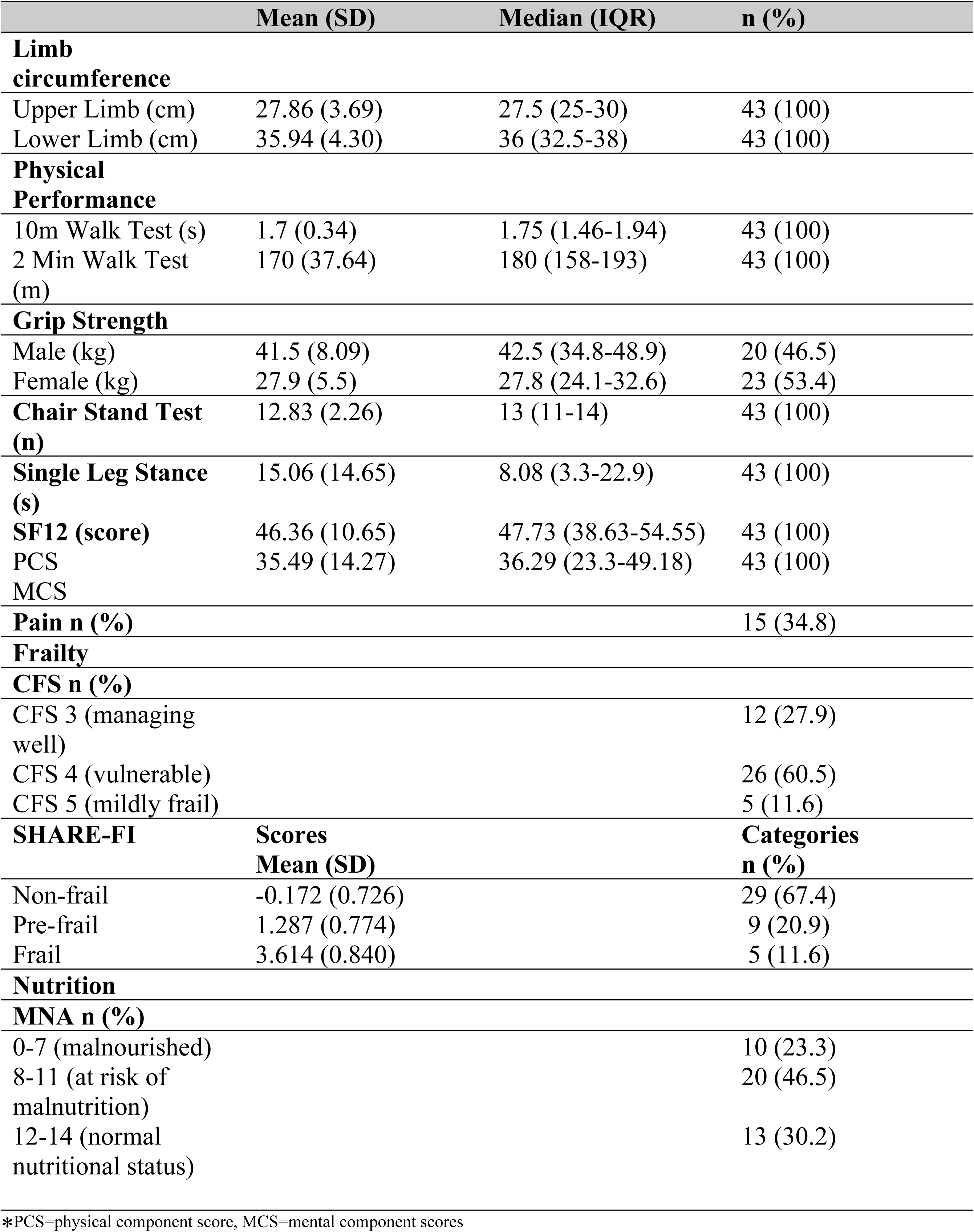
Baseline Physical Function and Frailty scores (n=43)

#### Retention

The overall retention rate was 69.8% with over two thirds (n=30) attending at least one session (Exercise Circuit or Park Walk). One quarter (25.6%, n=11) were *regular attenders*, under half (44.2%, n=19) were *sporadic attenders* and just under one third (30.2%, n=13) did not return following initial assessment (*non-attenders*) (Table I). Greater attendance was observed in the older participants with 72.7% (n=8/11) of the regular attenders over the median age (48 years), while 42.1% (8/19) of the sporadic attenders were over the median age. Greater female attendance was observed. Over one third (34.7% (n=8/23)) of females (regular attenders) versus 15% (n=3/20) of males attended regularly.

#### Adherence

Adherence to the prescribed exercises and the protein supplement was 93% and 90%, respectively. Pain (n=7, 23.3%), assault or accidental injury (n=2, 6.6%), external stresses (n=3, 10%), time (n=2, 6.6%), personal preference and lack of motivation (n=4, 13.3%) were the reported reasons for the inability to complete the prescribed exercises and taste was the cited reason for declining the protein drink.

#### Adverse Events

No adverse events occurred during the entire programme.

#### Acceptability

The exit survey, which took approximately five minutes, was completed by 20(47%) participants. Three themes emerged:

##### Barriers to retention

This theme was highlighted by:

> *“I should have kept going but I was in such a bad place that I didn’t want to feel happy.” (P1)*
>
> *“I was more interested in the drink”. (P5)*
>
> *“… just things in my addiction”. (P12)*
>
> *“There was a lot going on for me.” (P16)*

##### Psychological benefits

The exercise classes were reported to be fun, inclusive and sociable. Music was important and enjoyed particularly by the female participants.

> *“It was an enjoyable kind of exercise.” (P29)*
>
> *“I felt alive, I walked out tall, and I didn’t walk in like that … gave me something to look forward to.” (P1)*
>
> *“I felt great after it, I often feel down in the dumps, and it brings me back up … I’ve often a lot going on at home. I feel I leave it all behind me when I come … getting out mixing with people is great. (P40)*
>
> *“I got pain relief, friendship, energy, positivity” (P12)*.

The Park Walk was considered more feasible for some who felt unable for the exercise class.

> *“The Park Walk was just great, just doing the walk and the having the chat with the girls was good.” (P4)*

##### Abstinence

Another important finding was the reported reduction in substance use for some (n=8, 40%). Participants reported that it helped to postpone the use of substances until later in the day.

> “*I don’t smoke as much weed … I don’t smoke as much cigarettes” (P2)*
>
> *“It was keeping me away from the weed” (P15)*
>
> *“it was another thing stopping me from using” (P47)*

### Baseline measures

Eleven percent (n=5) of females and 11% (n=5) of males had ‘moderately-low’ or ‘severely-low’ lower limb (LL) circumference measures while 9% (n=4) of females and 13.9% (n=6) of males were below the cut-off values for upper limb (UL) circumference [29, 30]. Eighty-eight percent (n=38) of participants scored below the mean normative values for grip strength [27]. Just over half (55% and 53.4%) scored below the normative and average values for the 2MWT and the SLST, respectively [34, 35]. Pain was prevalent in 34.8% (n=15). Twenty-three percent (n=10) were malnourished and a further 46.5% (n=20) were at risk of malnutrition. The proportion of living with some degree of frailty was 33% and 73% using the SHARE-FI (pre-frail or frail) and CFS (4 or more), respectively. The SF-12 highlighted below average scores [mean (SD)=50(10)] for physical and mental component scores (PCS/MCS) for the collective group when compared to 1998 normative values [42]. More than half (58%, n=25) scored below the mean PCS value, while 81%, (n=35) scored below the mean MCS value.

### Impact

Impact was measured by observing pre-post intervention changes in the 20 participants who completed the final evaluation. Overall improvements were demonstrated in the CST (improvement of 2 ± 3.06 stands, [95% CI (0.568 to 3.432), *t*(19)=2.922, *p*=0.009], the 10mWT [0.25 ± 0.48m, 95% CI(0.025 to 0.47), *t*(19)= 2.328, *p*=0.031)] and the MNA (*Z*=-2.181, *p*=0.029) (Table 5).

**Table 5.** Secondary Outcomes at 12-week follow-up (n=20)

|  | Pre-intervention |  | Post Intervention |  | Intervention Effect |  |
| --- | --- | --- | --- | --- | --- | --- |
|  |  | Number (%) |  | Number (%) | Mean/median difference (95% CI) | P |
| **Chair stand test** mean (SD) | 12.65 (2.23) | 20 (100) | 14.65 (3.54) | 20 (100) | 2.00(0.57 to 3.43) | **\*0.009 a** |
| **Grip strength** mean (SD) | 31.8 (8.99) | 20(100) | 32.58 (11.86) | 20 (100) | 0.78 (-1.34 to 2.91) | 0.452 a |
| **10m Walk Test** mean (SD) | 1.60 (0.32) | 20 (100) | 1.85 (0.5) | 20 (100) | 0.25 (0.02 to 0.47) | **\*0.031 a** |
| **2min Walk Test** median (IQR) | 180 (166-189) | 19(95) | 194 (150-220) | 19 (95) | 34(13 to 61) | 0.387 b |
| **Single leg stance** median (IQR) | 8.2 (4-33) | 19(95) | 14 (6-36) | 19 (95) | 10.6 (2.80 to 23.50) | 0.355 b |
| **UL circumference** mean (SD) | 27.57 (3.88) | 20 (100) | 28 (4.07) | 20 (100) | 0.43 (-0.24 to 1.09) | 0.201 a |
| **LL circumference** mean (SD) | 36.14 (4.35) | 20 (100) | 36.52 (4.1) | 20 (100) | 0.38 (-0.25 to 1.01) | 0.218 a |
| **MNA** median (IQR) | 9.5 (6.25-12) | 20 (100) | 12 (9-12) | 20 (100) | 3 (0.00 to 4.00) | **\*0.029 b** |
| **Pain** median (IQR) | 3 (0-6.5) | 13 (65) | 0 (0-5) | 13 (65) | 0 (0.00 to 7.00) | 0.240 b |
| **SF12** |  |  |  |  |  |  |
| PCS median (IQR) | 44.7 (35.1-55.4) | 17 (85) | 48.9 (45.4-53.84) | 17 (85) | 10.7 (1.51 to 17.46) | 0.381 b |
| MCS | 38.61 (12.15) | 17(85) | 37.35 (12.44) | 17 (85) | 1.26 (-7.89 to 10.42) | 0.774 a |
| **Frailty** |  |  |  |  |  |  |
| **Clinical Frailty Scale** |  | 20(100) |  | 20(100) | 0(0.00 to 0.00) | 0.096 b |
| CFS 3 (Managing well) |  | 5 (25) |  | 9 (45) |  |  |
| CFS 4 (Vulnerable) |  | 13 (65) |  | 10 (50) |  |  |
| CFS 5 (Mildly frail) |  | 2 (10) |  | 1 (5) |  |  |
| **SHARE-FI** |  | 20(100) |  | 20(100) | 0(0.00 -0.00) | 0.248 b |
| Non-frail |  | 11 (55) |  | 14 (70) |  |  |
| Pre-frail |  | 6 (30) |  | 5 (25) |  |  |
| Frail |  | 3 (15) |  | 1 (5) |  |  |
| **SHARE-FI continuous scores** Mean (SD) | 0.968 (1.602) | 20 (100) | 0.372 (1.365) | 20 (100) | -0.595 (2.224) | 0.246 a |

a=paired samples t-test comparing pre- to post- intervention, b=Wilcoxin signed-rank test, \* = significant result; SF12 PCS = Physical Component Score, MCS = Mental Component Score

Data was stratified into regular and sporadic attenders. In the *regular attenders* (Table 6) improvements were observed in the 10mWT (0.428 ±0.48, 95% CI:(0.10 to 0.75); *t*(10)=2.943, *p*=0.015), the MNA (2.00 ±2.93, 95% CI(0.299 to 3.97); *t*(10) = 2.262, *p*=0.047) and in pain levels (*Z*=-2.214, *p*=0.027). Additionally, an improvement was detected in the CFS (*Z*= −2.121, *p*= 0.034) and SHARE-FI continuous score [95% CI (−2.805 to −0.73, *p=0.041].* Evidence was lacking for effectiveness in all outcomes for the *sporadic attenders*.

**Table 6:** Secondary Outcomes at 12-week follow-up (Regular Attenders)

|  | Pre-intervention |  | Post Intervention |  | Intervention Effect |  |
| --- | --- | --- | --- | --- | --- | --- |
|  |  | Number (%) |  | Number (%) | Mean/median difference (95% CI) | P |
| **Chair stand test** mean (SD) | 13.27 (2.19) | 11(55) | 15.45(3.29) | 11(55) | 2.18(-0.19 to 4.56) | 0.068 a |
| **Grip strength** mean (SD) | 29.07 (7.96) | 11(55) | 29.58 (10.09) | 11(55) | 0.51(-2.17 to 3.19)) | 0.682 a |
| **10m Walk Test** mean (SD) | 1.65 (0.29) | 11(55) | 2.07 (0.44) | 11(55) | 0.48 (0.10 to 0.75) | **\*0.015 a** |
| **2min Walk Test** mean (SD) | 180.90 (18) | 11(55) | 193.36 (32.1) | 11(55) | 12.45(-9.05 to 33.96) | 0.226 a |
| **Single leg stance** mean (SD) | 20.9 (15.30) | 11(55) | 25.56 (21.64) | 11(55) | 4.63(-7.2 to 16.46) | 0.404 a |
| **UL circumference** mean (SD) | 27. 90 (4.64) | 11(55) | 28.45 (4.62) | 11(55) | 0.55(-0.48 to 1.58) | 0.267 a |
| **LL circumference** mean (SD) | 36.20 (4.7) | 11(55) | 36.45 (4.57) | 11(55) | 0.25(-0.47 to 0.96)) | 0.464 a |
| **MNA** mean (SD) | 8.7 (3.13) | 11(55) | 10.7 (2.32) | 11(55) | 2.00(0.29 to 3.97) | **\*0.047 a** |
| **Pain** median (IQR) | 5 (0-6.5) | 9(45) | 0 (0-4) | 9(45) | 0 (0.00 to 0.00) | **\*0.027b** |
| **SF12** |  |  |  |  |  |  |
| PCS | 42.3 (10.4) | 11 (55) | 49.4 (6.8) | 11(55) | -3.7 (-2.81 to 16.99) | 0.142 |
| MCS | 37.0 (13.5) | 11 (55) | 39.5 (13.2) | 11(55) | 2.49 (-10.7 to 15.75) | 0.684 |
| **Frailty** |  |  |  |  |  |  |
| **Clinical Frailty Scale** |  | 11(55) |  | 11(55) | 0(0.00 to 00.00) | **\*0.034b** |
| CFS 3 (Managing well) |  | 4(36.4) |  | 8(72.7) |  |  |
| CFS 4 (Vulnerable) |  | 5(45.5) |  | 3(27.3) |  |  |
| CFS 5 (Mildly frail) |  | 2(18.2) |  | 0(0) |  |  |
| **SHARE-FI** |  | 11(55) |  | 11(55) | 0(0.00 to 0.00)) | 0.063 b |
| Non-frail |  | 6(54.5) |  | 9(81.8) |  |  |
| Pre-frail |  | 2(18.2) |  | 2(18.2) |  |  |
| Frail |  | 3(27.3) |  | 9(0) |  |  |
| **SHARE-FI continuous score** | 1.235 (1.904) | 11(55) | -0.204 (0.755) | 11(55) | -1.439 (2.033) | **\*0.041 a** |

a=paired samples t-test comparing pre- to post- intervention, b=Wilcoxin signed-rank test, \* = significant result; SF12 PCS = Physical Component Score, MCS = Mental Component Score

## Discussion

Key findings in this study indicate feasibility, evidenced by safety, acceptability, adherence, and retention in certain sub-groups. Impact was demonstrated in the regular attenders in pre-to post-intervention changes for a number of outcomes indicating proof of concept.

Initially to focus on retention, many did not return or came infrequently following the initial evaluation. It is posited that there are many reasons for the low retention observed. Survey feedback indicated the primary reason being chronic addiction challenges. Furthermore, those with unstable or inadequate accommodation attended less. The participants in this study lived in one of the highest areas of deprivation nationally within Ireland [46]. These findings were also demonstrated in the LEAP-1 trial, where stability in housing and addiction, appeared to drive better retention [19]. Furthermore, often observed was the presence of a tri-morbidity of substance misuse, a mental health disorder and a physical health condition. Many participants alluded to prior trauma, and it is suggested that adverse experiences, known to impact the life course likely influenced participants’ ability to engage. It is clear that the co-existence of these complex social factors contributed to the low retention observed.

In this study, health literacy appeared to be low in terms of realising the impact of risky lifestyle behaviours. Many participants smoked cigarettes prior to and immediately following exercise. Some participants declined to exercise due to reported health conditions and for them exercise was viewed as a futile activity rather than a valued opportunity to improve their health. Concurring with these findings, the literature has shown that over 90% of patients in receipt of addiction services demonstrate unhealthy lifestyle behaviours [47]. Moreover, lower health literacy levels in people with SUD have been linked to poorer quality of life and mental health [48]. These barriers combined with poor baseline physical activity levels were considered to impact negatively on motivation. Health promoting and awareness programmes preceding health interventions are proposed to enhance health literacy and retention.

Numerous strategies and behavioural change techniques to have been proposed to enhance adherence and outcomes in physical activity interventions, many of which were utilised in this study [49, 50, 51]. Among these are: exploring barriers and facilitators; improved participant knowledge and expectations, enjoyment and social support; goal setting and self-efficacy, and multi-disciplinary involvement [49, 50]. Finally, stakeholder involvement, also a feature in this study, particularly of those with lived experience of the condition under investigation is advocated in the planning stages of trials [52].

Findings of higher retention in the older and female participants in this study similarly emerged from the LEAP-I trial [19]. This may be due to a greater physical health need associated with ageing as well as the benefits of social engagement observed in the females. Another distinct group who engaged well were the female Travellers in this study. Feedback from these women who came from a nearby halting site (designated accommodation site for Travellers) was overwhelmingly positive. Factors influencing retention in this group were enjoyment and psychosocial support that the group offered, a cited distraction from difficult and often stressful living conditions. Irish Travellers, an indigenous Irish ethnic group, are known to suffer a disproportionate burden of physical health conditions compared to the general Irish population and their inclusion and engagement in this programme is considered important [53]. Furthermore, women experiencing homelessness and addiction have unique needs and this gendered dimension to the programme highlights a need for bespoke women-only services [54].

Inclusion of the Park Walk was important in this study which offered an alternative mode of exercise for some. Some preferred the “walk and talk” and enjoyed the “chat with the girls”. There is growing evidence of the restorative powers of exercising in open green spaces, improving cardiovascular markers as well as mental health outcomes [54–56]. Finally, other positive indicators in this study were high adherence to the exercises and the protein supplement as well as the absence of adverse events.

This study, in harmony with linked studies [3, 19] highlighted sub-normative values for physical functioning and frailty outcomes, validating the need for a targeted intervention. In terms of impact, sub-group analysis demonstrated that improvements were observed in the regular attenders only. The minimal clinically detectable improvement (MCID) for the CST in older adults is one stand and, in this study, the mean difference was two [58]. The MCID for the 10mWT in older adults is 0.05m/s [59] and in this study the change in gait speed was 0.25m/s. Movement components of the CST and brisk walking featured in the programme and were reportedly practiced outside of the programme and may have influenced improvements observed. Nutritional levels also improved. The post-exercise consumption of protein supplement regularly stimulated discussion around healthy eating and healthy behaviours which may have contributed to the enhanced nutritional scores detected.

Baseline levels of frailty in this study were relatively high (11% SHARE-FI and CFS). The SHARE study of older adults demonstrated frailty scores of 7.3% using the SHARE-FI [38]. Similarly, using the CFS, levels of frailty in The Irish Longitudinal Study of Ageing were lower (10.5%) than this study and the mean age was 20 years older (75.4 years) [60]. These outcomes evince the manifestation of accelerated ageing in this population and need for early intervention. An interesting finding was that the prevalence of living with some degree of frailty was much higher by CFS (73% ≥4) than the phenotypical SHARE-FI (33% pre-frail/frail), indicative of other drivers of frailty in this chronologically young population. Effectiveness of exercise interventions with nutritional supplementation to improve the frailty status in older adults has been demonstrated [22, 23]. In this study, an improvement was observed in the regular attenders using the CFS and in the continuous scores of the SHARE-FI. Nonetheless, post-intervention frailty (frail/pre-frail) using both measures (CFS and SHARE-categorical and continuous scores) was lower, and it is posited that even the postponement of decline, particularly in extremely vulnerable non-geriatric populations, is a positive outcome and such markers need to be acknowledged.

This study has addressed a research gap regarding frailty-focussed interventions in this population. It appears that this study, in harmony with the LEAP-1 trial, is one of the first to provide a structured exercise intervention to target frailty in this population. Its strengths are its pragmatic, unique low-threshold design, coupled with its’ co-design methodology. The exit survey highlighted important issues regarding retention and the impact exercise may have on abstinence and psychological wellbeing. Anecdotal feedback from stakeholders indicated that positive behaviours persisted beyond the lifecycle of the programme where a number of participants (n=6) continued the Park Walk and completed the Women’s 10km Mini Marathon two months later and others went on to engage in drug rehabilitation programmes. This concurs with the evidence where physical activity interventions translate into positive effects on wider life and addiction [20]. Mainstream services are unlikely to reach this population and low-threshold services are initially key to the delivery of interventions and may work as a bridge to mainstream services and more positive health behaviours.

### Limitations

Limitations of this study were the small sample size and absence of a control group. The authors also acknowledge that generalizability of its findings may be limited in general populations but may have important application to marginalised groups with addiction and homelessness challenges.

## Conclusion

This pragmatic study offers a valuable contribution to an emerging area and highlights the need for interventions to enhance physical health and delay the onset of frailty syndromes in this non-geriatric population. Greater consideration of the complex presentations of this population who face immense challenges to participation in health interventions is warranted. Further higher-powered research to evaluate effectiveness of exercise interventions and identify retention-enhancing strategies is warranted, particularly in sub-groups where better adherence and impact was observed. Nonetheless, where possible, all potential participants regardless of addiction issues and other challenges should be offered the opportunity to participate in exercise and physical rehabilitation programmes due to the breath of potential impacts demonstrated in this study and possibility of translation of these benefits to wider life. This research may be useful to drive policy in this area and guide frontline staff to deliver appropriate and flexible person-centred interventions for this hard-to-reach population.

## Data Availability

The data underlying this article cannot be shared publicly for the privacy of individuals that participated in this study. The data will be shared on reasonable request to the corresponding author.

## Acknowledgements

The authors wish to extend their gratitude to the study participants who committed their time to participate and the staff of the Ballyfermot Advance Project who assisted with this study.

## Supporting Information

**S1 Fig. PRISMA flow diagram of participants through the study**

**S2 Fig. Exit Survey**

